# Cerebral microbleeds in critically ill patients with respiratory failure or sepsis: a scoping review

**DOI:** 10.1101/2023.09.28.23296322

**Authors:** Bing Yu Chen, Johnny Dang, Sung-Min Cho, Mary Pat Harnegie, Ken Uchino

## Abstract

**Background:** Cerebral microbleeds (CMB) have been described in critically ill patients with respiratory failure, acute respiratory distress syndrome (ARDS), or sepsis. This scoping review aimed to systematically summarize existing literature on critical illness-associated CMBs.

**Methods:** Studies reporting on adults admitted to the intensive care unit for respiratory failure, ARDS or sepsis with evidence of CMBs on magnetic resonance imaging were included for review following a systematic search across five databases (MEDLINE, Embase, CENTRAL, Scopus, and Web of Science) and a two-stage screening process. Studies were excluded if patients’ CMBs were clearly explained by another process of neurological injury.

**Results:** Forty-eight studies reporting on 216 critically ill patients (mean age 57.9, 18.4% female) with CMBs were included. Of 216, 197 (91.2%) patients developed respiratory failure or ARDS, 5 (2.3%) developed sepsis, and 14 (6.5%) developed both respiratory failure and sepsis. Of 211 with respiratory failure, 160 (75.8%) patients had Coronavirus Disease 2019 (COVID-19). The prevalence of CMBs among critically ill patients with respiratory failure or ARDS was 30.0% (111 of 370 patients in cohort studies). The corpus callosum and juxtacortical area were the most frequently involved sites for CMBs (64.8% and 41.7% of all 216 patients, respectively).

Functional outcomes were only reported in 48 patients, among whom 31 (64.6%) were independent at discharge, 4 (8.3%) were dependent at discharge, and 13 (27.1%) did not survive until discharge. Cognitive outcomes were only reported in 11 of 216 patients (5.1%), all of which showed cognitive deficits (9 patients with executive dysfunction, 2 patients with memory deficits).

**Conclusions:** CMBs are commonly reported in patients with critical illness due to respiratory failure, ARDS, or sepsis. CMBs had a predilection for the corpus callosum and juxtacortical area, which may be specific to critical illness-associated CMBs. Functional and cognitive outcomes of these lesions are largely unknown.

## Introduction

Cerebral microbleeds (CMB) are accumulations of small hemosiderin deposits correlating with hypointense foci on susceptibility-weighted magnetic resonance imaging (MRI).^1^ CMBs are associated with several medical conditions, including cerebral amyloid angiopathy (CAA)^2^, traumatic brain injury (TBI)^3^ and chronic hypertension^4^. CMBs have been reported in critically ill patients among whom an etiologic explanation for cerebral microvascular injuries was not found.^5,6^ Common factors in these patient populations included the presence of respiratory failure or sepsis, possibly causing hypoxemia-induced microangiopathy or sepsis-related coagulopathy.^6^ The objective of this study was to conduct a scoping review describing available evidence on CMBs in critically ill patients with respiratory failure including acute respiratory distress syndrome (ARDS), or sepsis.

## Methods

### Design

This scoping review was conducted in accordance with recommendations from the Joanna Briggs Institute^7^ and the Preferred Reporting Items for Systematic Reviews and Meta-Analyses (PRISMA) extension for scoping reviews^8^ (Supplemental Table S1). As per these guidelines, a formal bias appraisal was not necessary and was not completed. Institutional review board approval was not required for conducting this review. Inclusion criteria were studies with patients age 18 or older, were critically ill due to respiratory failure, including ARDS, or sepsis, requiring admission to the intensive care unit (ICU), and showed evidence of CMBs on MRI. Studies were excluded if patients’ CMBs were clearly explained by another process of neurological injury, such as acute cerebral infarction, TBI, encephalitis, anoxic cerebral injury due to cardiac arrest, and infective endocarditis. All study designs were included except for reviews, guidelines and commentaries. Abstracts were included if results were not also published as manuscripts. Included studies must be published in English.

### Search strategy and selection

Five databases, MEDLINE (1946-), Embase (1974-), CENTRAL (1996-), Scopus (2000-), and Web of Science (1975-), were searched from their dates of inception until June 2, 2023. Key concepts related to microbleeds, critical illness, respiratory failure, ARDS, and sepsis were mapped to keywords and Medical Subject Headings terms. An example of search strategy used in MEDLINE is provided in Supplemental Table S2. Using a two-stage screening process, two co-authors (B.Y.C. and J.D.) independently selected relevant studies for subsequent data charting from a list of all studies identified using search strategies. In the first stage, studies were excluded based on abstract screening. In the second stage, studies were excluded based on full-text review. Screening was conducted using Covidence Systematic Review software. Any disagreement between the two co-authors was resolved by consensus.

### Data charting and synthesis

Two co-authors (B.Y.C. and J.D.) independently performed data charting. Final data charting was completed and verified by the first author. Synthesis and comparison of clinical, paraclinical, radiological and prognostic data were completed using representative tables and figures. CMB was defined according to the Greenberg’s criteria.^9^ ARDS was defined according to the Berlin definition.^10^ Disseminated intravascular coagulation was defined according to the International Society on Thrombosis and Haemostasis Criteria.^11^ Functional outcomes were trichotomized into independence (modified Rankin Scale or mRS 0-2), dependence (mRS 3-5), or death at the time of hospital discharge if outcome data from included studies permitted such characterization. Cognitive deficits during post-discharge follow-up assessment, if described in included studies, were classified into executive, memory, visuospatial, language, praxis or behavioral dysfunction.

## Results

In total, 48 studies were included for data charting and synthesis (Figure 1), comprising of 28 case reports (58 patients), 2 case-control studies (31 patients), 16 retrospective cohort studies (66 patients), and 2 prospective cohort studies (61 patients), published between 2012 and 2023 from 16 countries. Thirty-one studies described at least one patient diagnosed with Coronavirus Disease 2019 (COVID-19). A summary of all included studies is listed in Supplemental Table S3.

**Figure 1.**
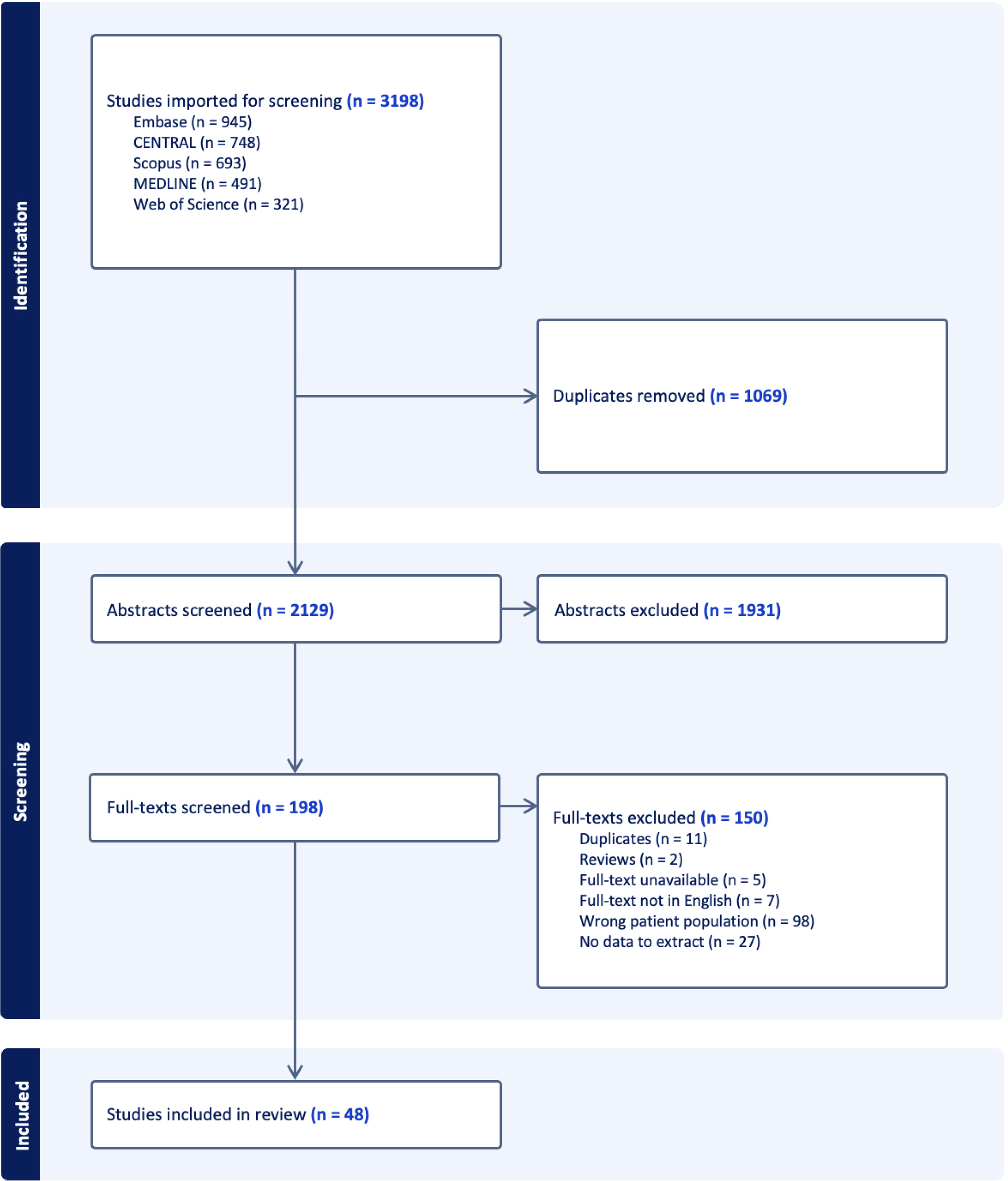
PRISMA flowchart of study selection.

A descriptive analysis of individual patient data extracted from included studies is shown in Table 1. A total of 216 critically ill patients with respiratory failure, ARDS or sepsis and findings of CMBs on MRI were included (mean age 57.9, 18.4% female). One hundred ninety-seven (91.2%) patients had developed respiratory failure or ARDS, 5 (2.3%) had developed sepsis, and 14 (6.5%) had developed both respiratory failure and sepsis, requiring ICU admission. One hundred sixty (74.1%) patients were diagnosed with COVID-19, and 172 (79.6%) were diagnosed with any form of pneumonia. During hospitalization, 114 (52.8%) patients required mechanical ventilation, 49 (22.7%) required extracorporeal membrane oxygenation (ECMO), 27 (12.5%) underwent dialysis, and 35 (16.2%) received therapeutic anticoagulation. Only 8 patients (3.7%) met diagnostic criteria for disseminated intravascular coagulation.

**Table 1.**
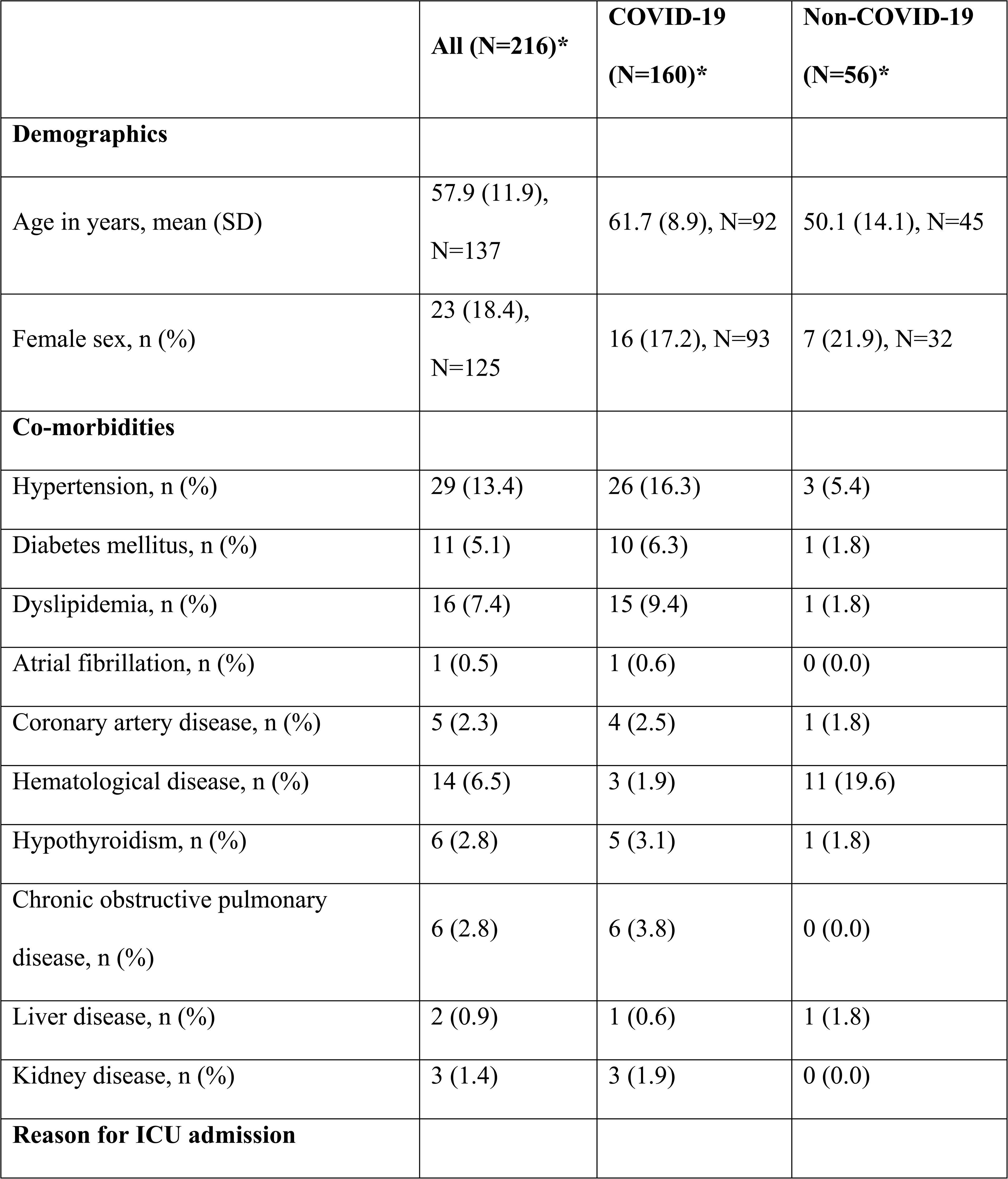
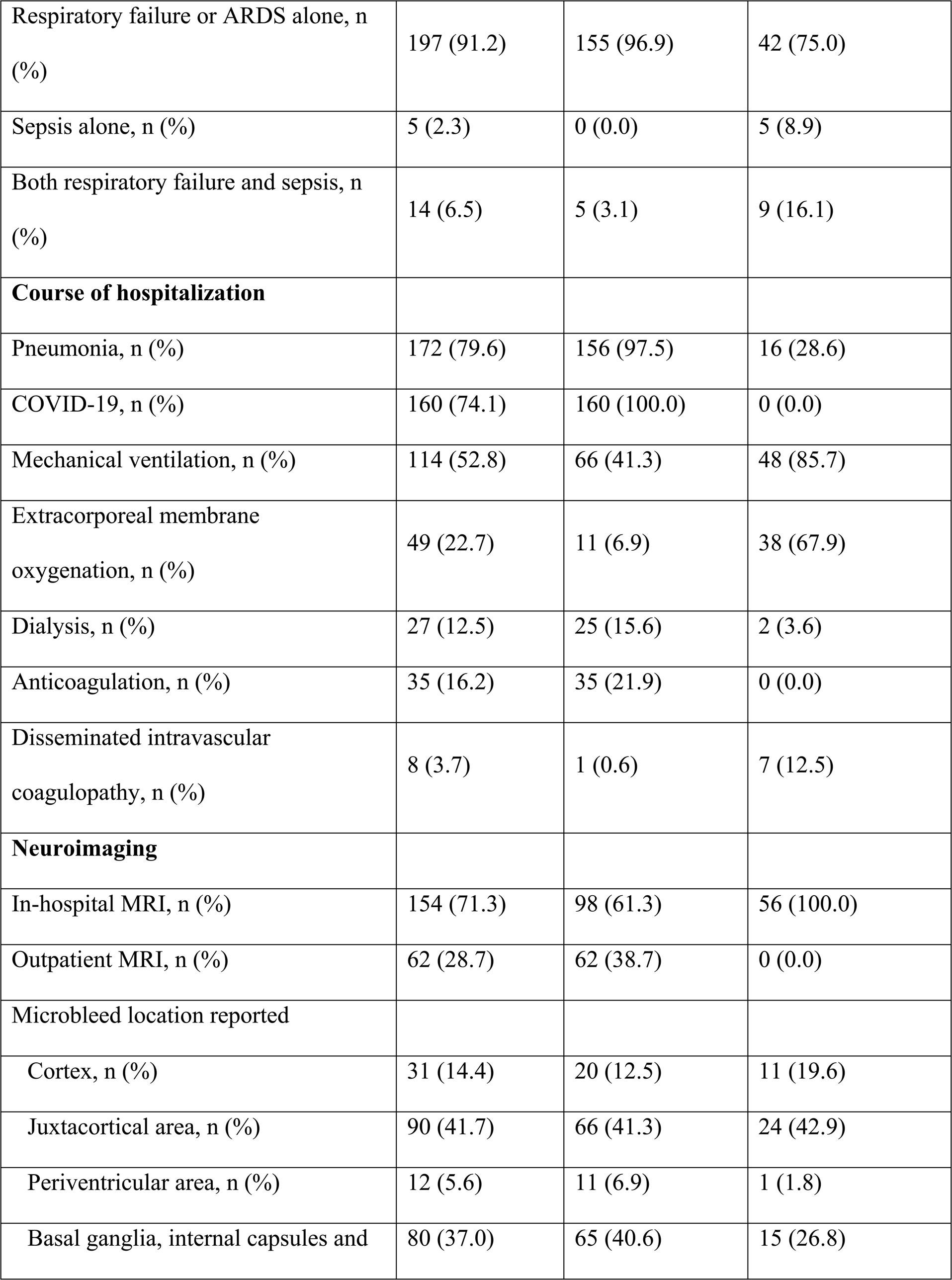
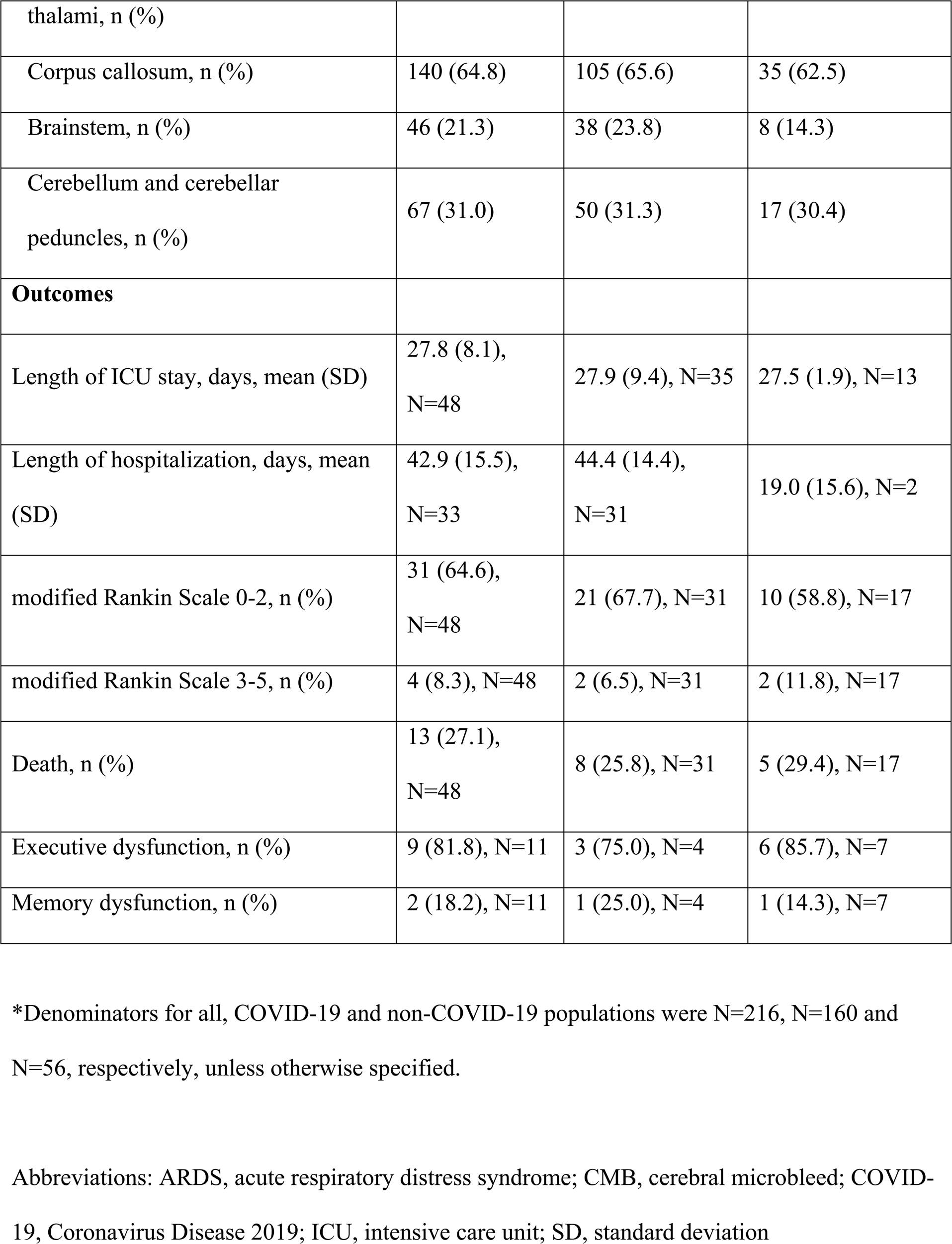
Patient characteristics, course of hospitalization, cerebral microbleeds location, and outcomes.

Of 7 studies providing data for estimating the prevalence of CMBs among critically ill patients with respiratory failure or ARDS undergoing MRI, the prevalence was 30.0% (111 of 370 patients in cohort studies, range 3.3% to 60.6%).^12–18^ The prevalence of CMBs among COVID-19 patients was 32.0% (99 of 309 patients, range 3.3% to 60.6%) and non-COVID-19 patients was 19.7% (12 of 61 patients, only one study reported prevalence).

One hundred fifty-four patients (71.3%) had at least one MRI scan of the head during hospitalization (median time 15 days after beginning of hospitalization, range 0-76 days, timing reported in 21 patients), while the remaining 62 (28.7%) only underwent MRI scan following discharge (median time 5 months after discharge, range 1-36 months, timing reported in 34 patients). The corpus callosum (140 of 216 patients, 64.8%) and juxtacortical area (90 of 216 patients, 41.7%) were the most frequently involved sites for CMBs (Figure 2). CMBs were often described as diffuse and innumerable by most studies, thus the number of CMBs in each location was not quantified in this study. In three studies, MRIs were performed both in-hospital and after hospital discharge, and in all 19 patients no change in the number and locations of CMBs was observed.^6,19,20^ Regardless of whether patients were diagnosed with COVID-19 or not, the most common sites of involvement for CMBs were the corpus callosum (105 of 160 (65.6%) COVID-19 patients, 35 of 56 (62.5%) non-COVID-19 patients) and juxtacortical area (66 of 160 (41.3%) COVID-19 patients, 24 of 56 (42.9%) non-COVID-19 patients).

**Figure 2.**
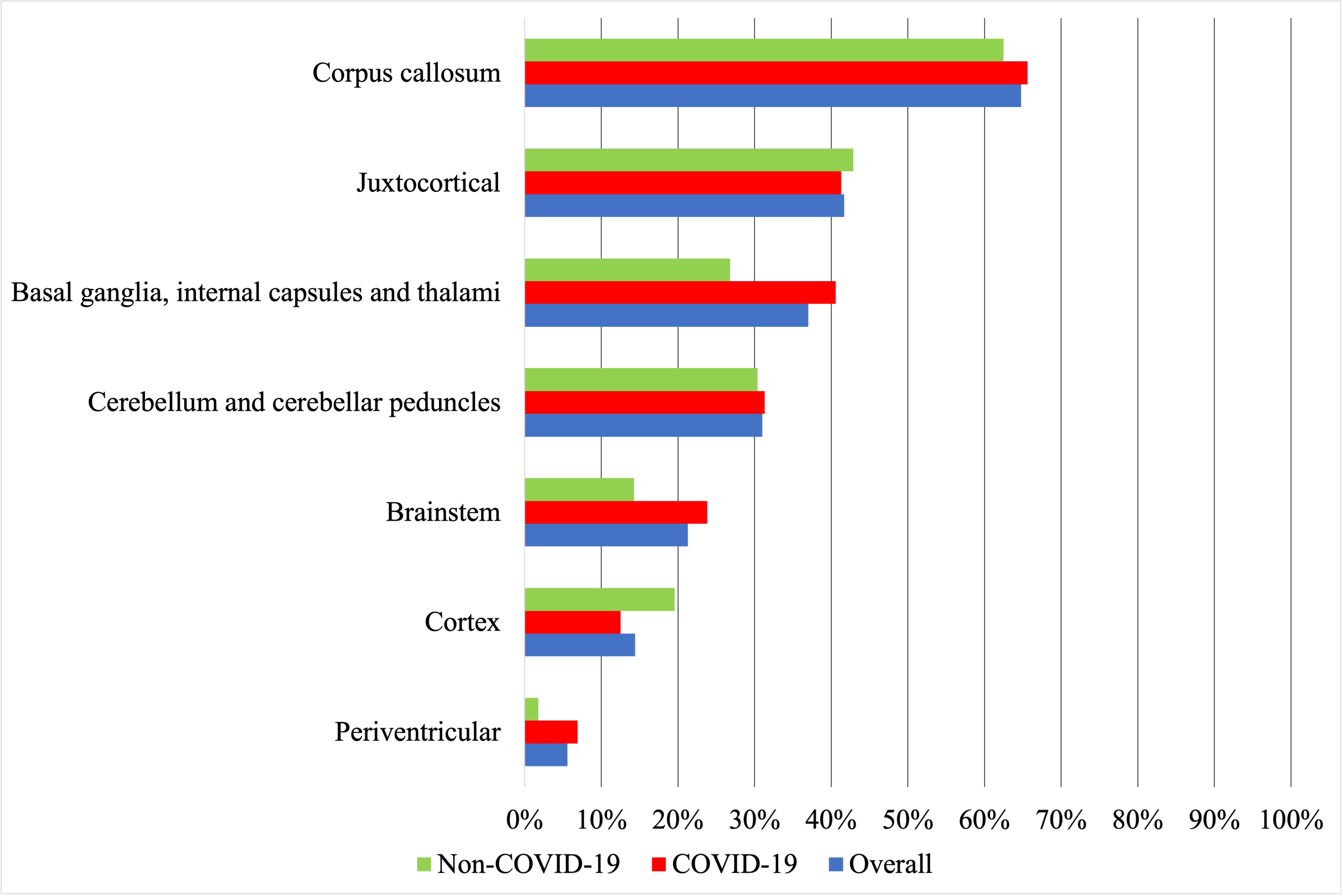
Cerebral microbleeds location.

Functional outcomes were only reported in 48 patients (22.2% of all patients with CMBs), among whom 31 (64.6%) were independent at discharge (mRS 0-2), 4 (8.3%) were dependent at discharge (mRS 3-5), and 13 (27.1%) did not survive until discharge. In the only study comparing outcomes between patients with and without CMBs, mortality at discharge was not different between the two groups (41.7% and 34.7% respectively, p=0.652).^18^ Cognitive assessment was reported in 11 patients (5.1% of all patients with CMBs) during follow-up (timing unavailable). All 11 patients had detectable cognitive deficits, 9 of which had executive deficits and 2 had memory deficits. No pre-hospitalization cognitive assessment was available for all 11 patients. No study comparing cognitive outcomes between patients with and without CMBs was found.

## Discussion

Our scoping review of CMBs on MRI among patients with critical illness due to respiratory failure, ARDS or sepsis resulted in 216 patients from 48 studies, comprised of 160 COVID-19 patients and 56 patients without COVID-19 infection. We found that the corpus callosum and juxtacortical area were the most common sites of involvement by CMBs. The prevalence and common sites of location and the CMB appear not to be different among the subgroup analysis of COVID-19 and non-COVID-19 cases, suggesting CMBs are likely reflective of respiratory failure, rather than COVID-19 infection.

The corpus callosum was the most common site, found in 64.8% of all patients, 65.6% of COVID-19 patients and 62.5% of non-COVID-19 patients. Callosal involvement by CMBs is commonly associated with TBI and generally not associated with other causes of CMBs (e.g. CAA, chronic hypertension, infective endocarditis, radiation).^21^ Patients with TBI were excluded from this review. CMBs in the corpus callosum have been described in patients with high-altitude cerebral edema (HACE).^22^ It has been hypothesized that severe hypoxemia in both HACE and critically ill patients may cause cerebral capillary vasodilation, disruption of the blood-brain barrier and extravasation of erythrocytes, particularly at the corpus callosum. Other hypothetical mechanisms for the presence of CMBs include disseminated intravascular coagulation, platelet dysfunction, and COVID-19 related endothelial inflammation.^23^ Iatrogenic causes of CMBs, such as ventilation-induced cerebral venous hypertension^5^, ECMO^24^, or dialysis^25^ have also been reported. In summary, hypoxemia and sepsis may result in disruption of the blood-brain barrier, coagulopathy, endothelial dysfunction, and CMBs.

Regardless of the pathophysiology, critical illness associated CMB could be considered a distinct entity given its unique preferential involvement of the corpus callosum and persistent CMB location pattern among both COVID-19 and non-COVID-19 patients. However, since only 2.3% of patients had isolated sepsis without respiratory failure or ARDS, it is difficult to establish whether patients with sepsis share a similar pattern of CMB location than patients with respiratory failure. Furthermore, the low number of published cases describing CMBs in patients with isolated sepsis may raise doubt on whether severe sepsis is strongly associated with CMBs. Critical illness associated CMB may be present mainly in respiratory failure or ARDS. This review only included patients with respiratory failure, ARDS or sepsis, mirroring a similar patient population described in the case series by Fanou and colleagues^6^ and review by Puy and colleagues.^21^ It is unknown whether other causes of critical illness, such as cardiogenic shock, are associated with CMBs.

CMBs have been associated with worse cognitive function in other contexts, but little is known about the clinical significance of the presence of CMBs among patients in the ICU.^26^ Only 48 patients out of 216 had any description of functional outcomes following discharge. Cognitive deficits were reported in only 5.1% of patients, which likely were underestimated due to underreporting and lack of follow-up. No comparative study of cognitive outcomes between critically ill patients with and without CMBs was found in our systematic review.

Our paper has several limitations. Most of the included studies were case reports, with potential reporting bias, but the majority of patients were from prospective or retrospective cohort studies. There was methodological heterogeneity across studies. The prevalence of associated patient characteristics, course of hospitalization, neuroimaging, and outcomes may be underestimated since some studies might not have reported the variable. For example, the prevalence of mechanical ventilation of 52.8% among critically ill persons with respiratory failure or sepsis is low. Well-designed prospective registries of critically ill patients are needed to inform on the true prevalence of CMBs in this patient population and the clinical and functional significance of CMBs, so that timely rehabilitation and appropriate services upon discharge may be provided.

In conclusion, a subset of critically ill patients with respiratory failure harbor CMBs with predilection for the corpus callosum and juxtacortical area in the brain. The true prevalence, clinical correlates, prognostic significance of these CMBs are important gaps in knowledge to be addressed in the future.

## Data Availability

All data pertaining to this manuscript are available upon request within 7 years of the date of publication.

## Acknowledgments

None

## Disclosures

BYC: Nothing to disclose

JD: Nothing to disclose

SMC: Relationship with Hyperfine (SAFE-MRI ECMO Study Principal Investigator)

MPH: Nothing to disclose

KU: Serves in data safety monitoring boards for clinical trials by Evaheart, Inc, and Genentech, Inc.

## Supplemental Material

Tables S1–S3

References 1–49

